# Impact of inspiratory vs expiratory incentive spirometry on obstructive, restrictive, and mixed ventilatory impairments: Study protocol Effect of incentive spirometry on respiratory impairments

**DOI:** 10.1101/2021.11.18.21266498

**Authors:** Eniola Awolola Oladejo, Sonill Maharaj Sooknunan

**Author notes:** **Corresponding Author:** Eniola Awolola Oladejo, Department of Physiotherapy, School of Health Sciences, College of Health Sciences, University of KwaZulu-Natal, Durban, South Africa, **Email:**, Contact Number. Authors’ Contributions (CRediT author statement) **Awolola Eniola:** Conceptualization, Methodology, Software, Data curation, Writing-Original draft preparation. **Maharaj Sonill:** Visualization, Investigation, Supervision, Software Validation, Writing-Reviewing and Editing. **Funding** Awolola Eniola Oladejo funds the study. The institution has no interest or role in the study’s design, writing the manuscript, collecting data, and analyzing data. No external funding is received from any source for the study.

## Abstract

**Background:** Respiratory impairments are adjusted reduction in pulmonary function that manifests with adverse effects on an individual’s state of health. The basis for identifying these impairments requires differentiation between normal and abnormal ventilatory capacity to avoid inaccurate and potentially harmful drug prescriptions and a mismatch of diagnoses. Incentive Spirometry (IS) is a technique designed to help patients achieve a pre-set volume of airflow; the volume is determined from predicted values or baseline measurements.

**Methods:** Fifty-four (54) patients with Obstructive, Restrictive, or Mixed type of respiratory impairments attending the respiratory clinic of Lagos State University Teaching Hospital, Ikeja (LASUTH) will be assessed for eligibility for the study. The study design will be a double-blind, randomized control trial with six intervention groups and three placebo control groups.

The study’s independent variable will be an Inspiratory Incentive Spirometry (IIS) and Expiratory Incentive Spirometry (EIS), while Obstructive, Restrictive, and Mixed respiratory impairments will serve as the dependent variable for the study. Baseline Pulmonary Function Test (PFT) will be assessed on entry into the study, and after every procedure, six minutes’ walk (6MWT) test and CAT assessment test will be assessed at every two (2) weeks interval of the study. Data will be analyzed using descriptive and inferential statistics of repeated ANOVA; P<0.05.

**Discussion:** The study outcome will determine the relationship between inspiratory and expiratory incentive spirometry on respiratory impairments and deduce the best fit device for the respective impairment.

**Trial Registration:** www.pactr.org: PACTR202005904039357

## Background

Respiratory diseases are frequently established spirometrically because other pathological confirmation methods are invasive and not routinely available; airflow-obstruction or restrictive-pattern collectively referred to as spirometric respiratory impairment are often detected through spirometry (1). Asthma and COPD are classified as airflow-obstruction, while diseases that involve the chest wall, respiratory muscles, pleura, or lung parenchyma are classified as restrictive-pattern (2). According to Fletcher (3), respiratory symptoms like dyspnea are reported in a quarter to a third of the adult population. They are associated with adverse outcomes like increased disability, and the risk of death is prevalent in aging populations and are associated with adverse outcomes.

Chronic obstructive pulmonary disease, often referred to as COPD, is a group of progressive lung diseases commonly associated with emphysema, chronic bronchitis, or both, which can lead to airway obstruction following the destruction of air sacs in emphysema and mucus build-up in the case of bronchitis (4, 5).

According to Similowski, Agusti (6), COPD is characterized by progressive, partially reversible airflow obstruction and lung hyperinflation with significant extrapulmonary (systemic) manifestations and comorbid conditions, contributing to the severity of the disease in individual patients. On the other hand, asthma is a disease affecting more than 300 million persons worldwide, with approximately 250,000 annual deaths. It is characterized by airway narrowing leading to reversible airflow obstruction in association with airway hyperresponsiveness (AHR) and airway inflammation (7, 8). The inhaled corticosteroid has become the primary treatment agent for asthma management in the last couple of decades (9). The diagnostic criteria that define spirometric respiratory impairment are often based on the Global Initiative for Obstructive Lung Disease (GOLD), the threshold of <0.70 for the spirometric ratio of forced expiratory volume in 1-second (FEV1) to forced vital capacity (FVC) frequently misclassifies normal spirometry as airflow-obstruction in otherwise asymptomatic never-smokers, starting at age 45-50 (2, 10-12). Normal aging leads to increased variability in spirometric performance, the GOLD threshold of 80% predicted for FVC, a criterion for establishing restrictive-pattern assumes equivalence to the lower limit of normal across the adult lifespan (13, 14)

Based on the controversies around spirometric classifications’ reliability, this study was conceived and designed to establish the relationships between spirometry classified respiratory impairments and Physiotherapy treatment procedure for obstructive, restrictive, and mixed respiratory impairments.

### Statement of the problem

Respiratory impairment is established in older persons and the aging population; it is pertinent to providers in primary health care, pulmonary medicine, and other specialties (2). Non-respiratory factors may also lead to respiratory symptoms and reduced mobility in older persons; the respiratory impairment must be identified with a high diagnostic relevance (2, 15, 16).

According to Pellegrino, Viegi (15), respiratory impairment is typically established by conducting a pulmonary function test using a spirometer and subsequently categorized as airflow limitation (e.g., COPD or asthma) or restrictive pattern (e.g., interstitial lung disease or heart failure). The criteria that define this airflow limitation are based on the Global Initiative for Obstructive Lung Disease (17)

Based on performance guidelines published by the American Thoracic and European Respiratory Societies (ATS/ERS), spirometry may be conveniently performed using a portable handheld device by instructing the participant to perform a series of forceful and complete exhalation maneuvers, starting from maximal inspiration with the breathing maneuvers generating two specific lung volumes, namely the FVC (an untimed lung volume) and FEV1 (a timed lung volume) (15).

Diseases may lead to a more significant reduction in the timed lung volume than the untimed lung volume, the FEV1/FVC is reduced and defines airflow limitations due to airways obstruction, while diseases that lead to comparable reductions in the timed and untimed lung volumes include those which affect the chest wall and respiratory muscles which will lead to a normal FEV1/FVC but reduced FVC (15)

In managing respiratory diseases, lack of awareness, knowledge and often delay in recognition of the disease is one of the reasons why primary care practitioners and other health care providers may incorrectly diagnose or manage the condition (18)

Pulmonary function test is a simple and accurate tool to assess airflow obstruction, patients forced expiratory volume in 1 s (FEV1)/forced vital capacity ratio is reduced, and FEV1 is reduced in COPD (Gold 2013). A reversibility testing differentiates COPD from asthma as in COPD patients do not show reversibility in airflow obstruction after administration of bronchodilators (19)

Although studies have shown the efficacy of incentive spirometry in the management of respiratory conditions, none has established the actual relationship between the inspiratory and expiratory type of incentive spirometry on the various types of respiratory impairments. This study is designed to answer the following questions.

1. What will be the effect of the Inspiratory and Expiratory type of incentive spirometry on pulmonary parameters, CAT score, and the six-minute walk test (6MWT) of patients with obstructive, restrictive, or mixed types of ventilatory impairments?
2. Will Inspiratory incentive spirometry affect the pulmonary parameters, CAT score, and six-minute walk test (6MWT) of patients with obstructive, restrictive, or mixed types of ventilatory impairments?
3. Will Expiratory incentive spirometry affect the pulmonary parameters, CAT score, and six-minute walk test (6MWT) of patients with obstructive, restrictive, or mixed types of ventilatory impairments?

### Aim of the study

#### Overall aim of the study

The study’s overall aim is to determine the effect of Inspiratory and Expiratory type of incentive spirometry on pulmonary parameters, CAT score, and the six-minute walk test (6MWT) of patients with obstructive, restrictive, or mixed types of ventilatory impairments?

### Specific Objectives

To determine

i. The effect of Inspiratory incentive spirometry on FEV1, FVC, FEV1/FVC, PEFR, CAT score, and 6MWT of patients with obstructive, restrictive, or mixed type of ventilatory impairments.
ii. The effect of Expiratory incentive spirometry on FEV1, FVC, FEV1/FVC, PEFR, CAT score, and 6MWT of obstructive, restrictive, or mixed type of ventilatory impairments.

### Hypotheses

#### Main Hypothesis

##### Ho

There will be no significant difference between the Inspiratory and Expiratory type of incentive spirometry on obstructive, restrictive, or mixed types of ventilatory impairments.

##### Ha

There will be a significant difference between the Inspiratory and Expiratory type of incentive spirometry on obstructive, restrictive, or mixed types of ventilatory impairments.

#### Sub Hypotheses

1. There will be no significant difference in the Pulmonary parameters, CAT score, and 6MWT of patients with obstructive, restrictive, or mixed types of ventilatory impairments before and after using inspiratory incentive spirometry in the study group.
2. There will be no significant difference in the Pulmonary parameters, CAT score, and 6MWT of patients with obstructive, restrictive, or mixed type of ventilatory impairments before and after using expiratory incentive spirometry in the study group
3. There will be no significant difference in the Pulmonary parameters, CAT score, and 6MWT of patients with obstructive, restrictive, or mixed types of ventilatory impairments before and after using placebo in the control group.

### Delimitation

This study will be delimited to 54 patients with COPD attending Lagos State University Teaching Hospital’s respiratory clinic, Ikeja, Lagos.

### Significance of the study

It is expected that the outcome of this study will establish the relationship between Inspiratory and expiratory type of incentive spirometry in the management of Obstructive, Restrictive, and Mixed type of respiratory impairments.

## METHOD

### Study Design

The study is a parallel 12-week randomized control trial. The study will involve three (3) groups; two intervention groups (IIS and EIS) and a placebo control group.

### Participants

The participants for this study will consist of male and female adult COPD patients aged 40 years and above attending Lagos State University Teaching Hospital (LASUTH), Ikeja, Lagos, Nigeria, West African.

*The inclusion criteria involve patients with a COPD history of about or more than six months duration attending the respiratory clinic of LASUTH*.

*The exclusion criteria involve patients with COPD on a cardiac pacemaker, supplemental oxygen therapy, asthma, cardiac conditions, and patients with psychological impairments*. Participants who met the required criteria will be asked to read and sign an informed consent approval for this study by the appropriate institutional review board.

### Setting

Patients with COPD attending the respiratory clinic of Lagos State University Teaching Hospital Ikeja, Lagos State, Southwest Nigeria, would be recruited for the study. The hospital is a tertiary health care facility in Lagos state and receives referrals from within and outside the state.

#### Sample size

Pulmonary function test is the primary outcome of interest for the study and the expected clinically relevant difference for obstructive restrictive and mixed impairment using LLN and GLI reference equation proposed by Quanjer et al. (2012). Therefore, the sample size (N) will be determined using G-Power statistics software. The power is selected at 95%=0.95, confidence level at 5% =0.05 and effect size of 0.35. We intend to investigate the main effects, within and between interactions for two factors, namely:

Factor A – Therapy with three levels (Therapy A, Therapy B, and No Therapy)

Factor B – Types of impairments (Obstructive, Restrictive and Mixed), nested within factor A

This gives 3×3 = 9 groups of subjects from which repeated measurements are to be obtained. We are also planning to measure three continuous outcomes at different time points, of which the minimum repeated measurements are expected to be 6 per subject for some of the outcomes and a maximum of 36 per subject. If a minimum of 6 repeated measurements is to be obtained from each subject, it is estimated that a minimum total sample size of 51 patients is required to detect an effect size v = 0.35 with 95% confidence (5% Type I) and having similar studies also expected to produce similar results about 80% of the time (Power of test 80%). In the case that during the time of the data collection, the hospital experiences a heavy burden of COPD patients or resources permit, there is an allowance to increase the total sample size to at most 68 subjects with a corresponding increase in the power of the test to about 95% and still detecting effect sizes between 0.3 and 0.4 (See Figure 1). This translates to the possibility of recruiting between 5 and 8 patients per group. Hence, the sample size estimates suggest that recruiting more than eight patients per group for this kind of study will be a waste of resources.

### Randomization and blinding

The contact numbers of participants will be randomly extracted from the respiratory patients’ database attending Lagos State University Teaching Hospital Ikeja’s respiratory clinic. A bulk Text message captioned “Invitation to a study on respiratory impairments will be circulated using the Luxury bulk SMS platform. Respondents will be assessed for eligibility, and those that meet the inclusion and exclusion criteria will participate in the study.

Participants will be randomly selected by simple randomization using a computer software program (Suresh, 2011). We will use the software program www.randomization.com to assign interested participants into three major groups (groups A, B, and C); subgroup A (x, y,z), subgroup B (e, f,g), and subgroup C (h, i,j). Group A and B will represent the study group, while group C represents the control group. Subgroup x, e, and h represent patients with obstructive respiratory Impairments, Subgroup y, f and i represents patients with restrictive respiratory impairments, while Subgroup z, g and j connotes patients with mixed respiratory impairments.

#### Inspiratory Incentive Spirometry

Eighteen subjects will be assigned to any of the three subgroups labeled ‘x’ ‘y’ or ‘z,’ at the ratio of six subjects per subgroup, depending on their nature of impairments. The participants will be required to breathe in the upright position before commencing the exercise using a respiration method that is best convenient for them. After that, a volume-incentive spirometer (Coach 2 device), which enables the patient to inhale air through a mouthpiece, and corrugated tubing attached to a plastic bellows will be used for inspiratory exercise with the volume of air displaced indicated on a scale located on the device enclosure. After the patient has achieved the maximum volume, they are instructed to hold this volume constant for 3 to 5 seconds (20). A total of three sets will be performed up to a maximum of 10 attempts. One minute of rest will be given between sets to reduce muscular fatigue. The procedure will be repeated three times a week for 12 weeks. Pulmonary function test will be conducted before and after every procedure while the CAT score, Six-minute walk test will be conducted fortnightly.

#### Expiratory Incentive spirometer

Eighteen subjects will be assigned to any of the three subgroups labeled ‘e’ ‘f’ or ‘g,’ at the ratio of six subjects per subgroup, depending on their nature of impairments. The participants will be required to breathe in the upright position before commencing the exercise using a respiration method that is best convenient for them. They will be asked to breathe in air through the nose to the maximum effort, then breathe out into the balloon at the maximum rate and maintain this state for one second (20). They will be required to close the mouth of the balloon with their fingers immediately, breathe-in to the maximum once again, and then breathe out into the balloon. The balloon-blowing exercise will be performed two times at maximum balloon-blowing over one minute. The exercise will be repeated three times to complete one set; a total of three sets will be performed up to a maximum of 10 attempts. One minute of rest will be given between sets to reduce muscular fatigue. The procedure will be stopped whenever the subjects felt dizziness or experience pain in the chest. To avoid Valsalva during balloon-blowing, the subjects will be asked not to hold their breath for more than five seconds after the maximum expiration. The participant will be allowed to go home after the musculoskeletal assessment. The procedure will be repeated three times a week for 12 weeks. CAT score, Six minutes’ walk test will be conducted fortnightly.

#### Control group

Eighteen subjects will be assigned to any of the three subgroups labeled ‘h’ ‘i’ or ‘j,’ at the ratio of six subjects per subgroup, depending on their nature of impairments. Following a successful allocation, the participants will be required to breathe in the upright using a respiration method that is best convenient for them. An assessment of participants’ musculoskeletal system will be conducted immediately after his/her pulmonary function test and the result of the test will be explained to the patient. The participant will be allowed to go home after the musculoskeletal assessment. The procedure will be repeated three times a week for 12 weeks. CAT score and Six minute’s walk test will be conducted fortnightly.

### Outcome Measure

#### Spirometry Assessment

The assessment will be conducted by a spirometrist certified by the Pan African Thoracic Society. A portable spirometer (Koko SX 1000 Standalone Version 7 Pneumotach) will be used to carry out this assessment. Daily calibration of the device will be conducted using a 3.0-liter syringe. A brief description of the assessment procedure, including technical steps to obtain pulmonary function data and variables, will be explained to each subject. After 2-3 tidal breaths, subjects will be asked to inhale deeply to total lung capacity and then exhale rapidly (without any pause) through a disposable mouthpiece until as much air as possible has been expelled from the lungs. The test will be performed in a sitting or standing position. The assessments will be repeated three times after adequate rest. The maximum number of attempts permitted will be 8; after fulfilling the acceptability and repeatability criteria, the two best curves and two best tests will be selected. The average values of the forced vital capacity (FVC) and forced expiratory volume in the first second (FEV1) will be recorded (21)

#### Six-Minute Walk Test (6MWT)

The 6MWT will be performed according to the standardized procedure which the researcher will supervise. The patients will be asked to walk at their own maximal pace along a 30m long, flat, and straight hospital corridor. The patient will not be given any encouragement. Patients’ symptoms may limit the test; therefore, if any signs of significant distress such as dyspnea, dizziness, angina, or skeletal muscle pain, the patient will be asked to stop. The patient will only be allowed to continue upon abatement of the discomfort. The total distance covered by the patient will be recorded in meters.

#### CAT Score

According to Jones P.W. Jones (22), the CAT was created using COPD patients’ input, developed using modern questionnaire methodology, psychometric analysis, and item response theory using Rasch analysis to identify items with the best-fit from a unidimensional instrument. It is a self-administered questionnaire consists of eight items assessing various manifestations of COPD (cough, sputum, dyspnea, chest tightness) aiming to provide a simple quantifiable measure of HRQoL

CAT scores from 0–40, higher scores denote a more severe impact of COPD on a patient’s life, the difference between stable and exacerbation patients is five units, no target score represents the best achievable outcome, its intra-class correlation coefficient=0.8, its internal consistency Cronbach’s α=0.88 and has a high correlation with SGRQ (r=0.84) across 7 European countries (22).

### Data analysis

The Statistical Package for Social Sciences (SPSS Inc, Chicago, II) version 26.0 for the Windows package program will be used to analyze data. Descriptive statistics of mean, standard deviation, frequency, and percentage will be used to summarize the results. Bar charts, pie charts, and histograms will be utilized for pictorial illustration. A multilevel analysis of variance (ANOVA) will be used to compare the outcome pulmonary function variables (FEV1, FVC, FEV1/FVC, PEFR, CAT score, and 6MWT) among each group. A dependent t-test will be used to compare the pre and post-test results, while an independent t-test will be used to compare the outcome variable across the three groups. The level of significance will be set at p0.05.

### Harms

The research does not potentially involve risk or harm as all interventions can only be performed within the limit of the patient’s tolerance. However, those participants may experience mild discomfort due to the exercise, and this may include temporary muscle soreness, increase in heart rate, blood pressure, sweating, and dizziness. Although all necessary care will be in place to prevent the occurrence of any adverse event, however, in case of a report of serious adverse events (e.g., comorbidities, injuries, persistent excruciating pain, dizzy spells, headache) after intervention or at any point during the trial, then we would consider unblinding the participant to the intervention for his/her safety. Additionally, the participants will be instructed to report any adverse events to the PI or the physiotherapist supervising their group. The number of participants per group will be limited to a maximum of 3 per day to ensure the participants’ adequate supervision and safety. Arrangements have been made with the Accident and Emergency units, the hospital where the research will be conducted to provide a standby medical team. However, the University of KwaZulu-Natal insurance scheme on clinical trials has fully covered the participants in this type of study.

## Discussion

Despite a good number of randomized control trials on the effectiveness of incentive spirometry in managing respiratory impairments, there is a dearth of literature to identify the specific type of incentive spirometer most suitable for the obstructive, restrictive, and mixed type of respiratory impairments, respectively.

According to (23), Incentive Spirometry (IS) is a lung expansion technique designed to induce sighing or yawning by making the patient take long, slow deep breaths that prevent and treats inactive atelectasis patients who have a predisposition for shallow breathing.

Studies by Kumar, Alaparthi (23) on the effect of incentive spirometry on the patient who had undergone laparoscopic abdominal surgery, the inspiratory incentive spirometry resulted in early recovery of pulmonary function and diaphragm movement in patients.

Adeniyi and Saminu (24), on the effect of local incentive, spirometry on Patients with sickle cell anemia, a locally revealed an increase in the peak expiratory flow rate of the patients, similarly in a study by Kim and Lee (20), on the effect of balloon blowing exercise on lung function of young adult smokers revealed a significant improvement in Vital capacity, expiratory reserve volume, inspiratory reserve volume, forced vital capacity and forced expiratory volume in one second. Although studies by Kim and Lee (20), Adeniyi, and Saminu (24) identified the effect of inspiratory and expiratory type of incentive spirometry on sickle cell anemia and a population of smokers, its effect relative to the nature of impairments was not ascertained. Consequently, it is expected that the study’s outcome will help to understand further the effect of IIS and EIS on obstructive, restrictive, and mixed types of respiratory impairments.

Finally, it is expected that the findings of this study will be recommended in the clinical guidelines for the management of respiratory impairments and will support the cost-benefit of COPD management in Nigeria and other low-income countries.

### Access to Protocol

The protocol was registered on the 17th of May 2020 with identifier number PACTR202005904039357, and the trial organization is PACTR

## Data Availability

All data would be collected on a sheet of papers (form) and kept confidential under lock & key and transfer to excel sheet electronically on a password protected computer at the Physiotherapy department, college of health sciences, University of KwaZulu-Natal, Durban for 5 years, only the researchers will have access to it after this period all documentation will be destroyed.

## Acknowledgments

We want to thank all the physiotherapists and medical doctors for their contribution to the study’s success and, most importantly, the participants for their voluntary participation. Our profound appreciation goes to Dr. Olufunke Adeyeye and Dr. Olufemi Ojo of Lagos State University Teaching Hospital, Ikeja, Lagos, for their professional advice in writing and reviewing this manuscript, also Miss Kemi and Miss Laide for their support in Pulmonary function test, and Miss Amodeni Ayomopewa for her editorial input.

## Ethical Considerations and consent to participate

Biomedical Research Ethics Committee of the University of KwaZulu Natal (South Africa) approved this study with approval number BREC/00001883/2020, and the Human Research Ethics Committee of Lagos State University Teaching Hospital, Ikeja, Lagos, Nigeria, West Africa, approved the study site location with approval number LREC/O6/10/1455. The study has been registered with ClinicalTrial.gov with the following registration number: PACTR202005904039357. All participants for this study will be required to sign a written informed consent, the consent form will be administered by a third party who is independent of study team.

The consent form was designed by the Biomedical Research Ethics Committee of the University of KwaZulu-Natal according to the world medical association Helsinki declaration and good clinical practice (GCP).

The primary investigator will communicate in writing to the RECs in the event of the need to modify the protocol, especially inclusion or exclusion criteria of the study during the trial..

## Consent to publication

Not Applicable

## Availability of data and materials

The datasets used and analyzed during the current study are available from the corresponding author. However, the findings from the study would be made available to participating researchers as required by law.

## Abbreviations

6MWT: Six Minutes’ Walk Test
ANOVA: Analysis of Variance
CAT: COPD Assessment Test
COPD: Chronic Obstructive Pulmonary Disease
EIS: Expiratory Incentive Spirometry
FEV1: Forced Expiratory Volume on 1 Second.
FRC: Functional Residual Capacity
FVC: Forced Vital Capacity
GLI: Global lung Initiative.
IIS: Inspiratory Incentive Spirometry
LASUTH: Lagos State University Teaching Hospital
LLN: Lower Limit of Normal
MANOVA: Multiple Analyses of Variance
MS: Microsoft
PFT: Pulmonary Function Test
RCT: Randomized Control Trial
ROM: Range of Motion
SPSS: Statistical Package for Social Sciences
UKZN: University of Kwazulu-Natal

